# Sex-Specific Correlates of Schistosoma haematobium Infection in a Rural Zambian Setting: A pilot study Focusing on Immune and Hematological Factors

**DOI:** 10.64898/2025.12.08.25341872

**Authors:** Reggison Phiri, Emmanuel L. Luwaya, David Chisompola, Martin Chakulya, Lukundo Siame, Bislom C. Mweene, Benson M. Hamooya, Joseph M. Chalwe, Joreen P. Povia, Sepiso K. Masenga

## Abstract

**Background:** This study aimed to identify and compare the sex-specific immune, hematological, and environmental factors associated with *Schistosoma haematobium* infection in a rural Zambian setting.

**Methods:** This community-based cross-sectional study enrolled participants from rural Zambia. Schistosoma haematobium infection was diagnosed microscopically from urine. Blood samples were analyzed for complete blood counts and cytokines via ELISA. Data were analyzed using sex-stratified logistic regression models in StatCunch.

**Results:** The analysis included 51 male and 92 female participants. In the multivariate model, for males, only lack of clean water (aOR=0.04, 95% CI; 0.00-0.60, p=0.0194) and symptomatic disease (aOR=68.10, 95% CI; 4.64-1000.29, p=0.0021) remained independent correlates of schistosomiasis infection. For females, the final model identified symptomatic disease (aOR=2155.53, 95% CI; 2.11-2198049.30, p=0.0299), elevated IL-4 (aOR=1.02, 95% CI; 1.01-1.03, p=0.0035), lower hemoglobin (aOR=0.17, 95% CI; 0.03-0.87, p=0.0336), and a lower platelet-to-lymphocyte ratio (aOR=0.97, 95% CI; 0.93-1.00, p=0.0489) as significant independent correlates.

**Conclusion:** The correlates of schistosomiasis infection are highly sex specific. While environmental exposure is a key driver for males, the female profile is characterized by a pronounced immunological response, including a Th2-skewed cytokine profile and associated hematological manifestations like anemia. These findings underscore the necessity for sex-differentiated approaches in both the clinical assessment and public health control of schistosomiasis.

## Introduction

Schistosomiasis remains one of the most prevalent and devastating neglected tropical diseases (NTDs), with a global burden disproportionately shouldered by sub-Saharan Africa [1]. Of the human-pathogenic species, *Schistosoma haematobium* is responsible for urogenital schistosomiasis, a chronic infection estimated to affect over 110 million people, with the vast majority of cases concentrated in low income countries [2].

The pathology of *S. haematobium* arises primarily from parasite eggs becoming trapped in the walls of the bladder, ureters, and genital tissues. This triggers a granulomatous inflammatory response that can cause hematuria, fibrosis, urinary obstruction, and an increased risk of bladder cancer [3]. The host counters egg deposition with a T-helper 2 (Th2) immune response, characterized by the production of interleukin (IL)-4, IL-5, IL-13, and IgE [4,5]. While this Th2 response is essential for containing the infection, it is concurrently regulated by immunomodulatory pathways, such as regulatory T cells (Tregs) and IL-10, to prevent excessive tissue damage [6]. The important balance between these pro-inflammatory and regulatory forces ultimately determines infection outcomes, ranging from resistance to reinfection to the development of severe morbidity [7].

Biological sex is an important modifier of infection outcomes. Sex hormones directly influence immune function, and immunological dimorphism may explain sex-biased infection and pathology beyond just behavioral differences [8–13]. While murine models of *S. mansoni* show females mount stronger immune responses [14], sex-specific immune and hematological correlates in *S. haematobium* infection remain a significant knowledge gap. Furthermore, a growing body of evidence has established a robust association between female genital schistosomiasis (FGS) and heightened susceptibility to HIV, underscoring the infection’s role as a critical, yet often overlooked, co-morbidity [15–17].

In schistosomiasis, some studies have reported higher prevalence and infection intensities in males, often attributed to gender-based differences in water-contact behavior [18]. However, a purely behavioral explanation is insufficient, as emerging evidence points to intrinsic biological factors that may render one sex more susceptible to infection or pathology. Most immunopathological studies of schistosomiasis either do not stratify their analyses by sex or lack the power to detect sex-specific effects [19].

This omission is essential, as sex-specific biology may influence reinfection rates, pathology, and the success of public health interventions like mass drug administration (MDA) with praziquantel [20–22]. This study aims to identify the sex-specific immune and hematological correlates of S. haematobium infection in a rural Zambian community. We hypothesize that distinct profiles between males and females are linked to differences in infection intensity and morbidity, knowledge essential for developing more effective, stratified control strategies.

## Methodology

### Study Design

A community-based cross-sectional study was carried out in the rural communities of Machile, Sichili, and Mulobezi within the Mulobezi District of Western Zambia [23]. A total of 143 participants (92 females, 51 males) across pediatric and adult age groups were enrolled.

### Eligibility Criteria

The study enrolled residents of the district, aged 5 to 55 years, who had resided in the area for a minimum of three months. All participants provided informed consent; for children under 16 years, assent was obtained from the child alongside consent from a parent or guardian. Pregnant individuals and those with serious comorbidities, such as advanced HIV or autoimmune diseases, were excluded.

### Data and Sample Collection

Participants were recruited between 2^nd^ January 2026 and 1^st^ June 2024. Participants underwent a structured interview to gather demographic and behavioral data. Biological samples collected included two venous blood samples and a single midstream urine sample. The urine specimens were transported under cool conditions to a local hospital for immediate microscopic analysis. Blood samples were processed for a complete blood count at a certified clinical laboratory, while separated serum was shipped to a specialized research facility for cytokine quantification.

### Laboratory Procedures

Diagnosis of *S. haematobium* infection was confirmed through microscopic identification of ova in centrifuged urine sediment by two trained laboratory technologists. In case of disagreement a third laboratory technologist was involved to decide. Plasma concentrations of the cytokines IL-4 and IL-10 were determined using commercially available, validated enzyme-linked immunosorbent assay (ELISA) kits, adhering strictly to the manufacturer’s protocols.

### Statistical Analysis

Data were analyzed using StatCrunch software. Descriptive statistics summarized participant characteristics. The Mann-Whitney U test was employed for non-normally distributed continuous variables, and the Chi-square test assessed associations between categorical variables. To identify infection correlates, sex-stratified univariate and multivariable logistic regression models were constructed, with statistical significance defined as a p-value of less than 0.05.

### Ethical Considerations

The study protocol received full approval from Mulungushi University School of Medicine and Health Sciences Ethics Review Committee (Ref. No.: SMHS-MU4-2023-031) on 26^th^ December 2023. Prior to enrollment, all participants provided written informed consent/assent after a detailed explanation of the study in their local language, ensuring voluntary participation and confidentiality. Data were anonymized to protect confidentiality. All data collected and analyzed were de-identified to ensure complete confidentiality. No information leading to identification of patients during and after analysis was abstracted and entered in the data collection form.

## Results

### Characteristic of the study population

Baseline characteristics of the 143 participants, stratified by biological sex. Data are shown as median (IQR) or n (%). Statistical comparisons revealed that the male (n=51) and female (n=92) cohorts were well-matched, showing no significant differences in key exposure, clinical, or hematological parameters. The frequency of using protective measures was the only characteristic that differed significantly between the groups (p=0.011).

**Table 1.** Comparison of Participant Characteristics by Sex.

| Variable | Median (IQR)<br>/Frequency (%) | Male=51<br>(35.66) | Female=92<br>(64.34) | P value |
| --- | --- | --- | --- | --- |
| Age (yrs) | 15 (9,30) | 13 (8,20) | 16 (10.5,30) | 0.0632 |
| Access to clean water |  |  |  | 0.3992 |
| Yes | 55 (38,5) | 22 (40) | 33 (60) |  |
| No | 88 (61.5) | 29 (32.95) | 59 (67.05) |  |
| Source of drinking water |  |  |  |  |
| Tap | 55 (38.5) | 22 (40) | 32 (60) | 0.4862 |
| River | 88(61.5) | 29 (35.95) | 59 (67.05) |  |
| Signs/Symptoms |  |  |  | 0.4451 |
| Yes | 64 (44.8) | 25<br>(39.06) | 39<br>(60.94) |  |
| No | 79 (55.2) | 26<br>(32.91) | 53<br>(67.09) |  |
| *Schistosomia* |  |  |  |  |
| Yes | 56 (39.2) | 18<br>(32.14) | 38<br>(67.86) | 0.4806 |
| No | 87 (60.8) | 33<br>(37.93) | 54<br>(62.07) |  |
| *Have you ever*<br>*been diagnosed* |  |  |  | 0.6183 |
| Yes | 25 (17.5) | 10<br>(40) | 15<br>(60) |  |
| No | 118 (82.5) | 41<br>(34.75) | 77<br>(65.25) |  |
| *Ever been tested*<br>*for other parasitic* |  |  |  |  |
| Yes | 138 (96.5) | 2<br>(20) | 84<br>(63.16) | 0.495 |
| No | 5 (3.5) | 49<br>(36.84) | 8<br>(80) |  |
| *On treatment for*<br>*other condition* |  |  |  | 0.4167 |
| Yes | 16 (11.2) | 4<br>(25) | 80<br>(62.99) |  |
| No | 127 (88.8) | 47<br>(37.01) | 80<br>(62.99) |  |
| *Education* |  |  |  | 0.0553 |
| *Not Schooled* | 22 (15.4) | 7<br>(31.82) | 15<br>(68.18) |  |
| *Primary* | 102 (71.3) | 41<br>(40.2) | 61<br>(59.8) |  |
| *Secondary* | 18 (12.6) | 2<br>(11.11) | 16<br>(88.89) |  |
| *Tertiary* | 1 (0.7) | 1<br>(100) | 0 |  |
| *Protective*<br>*measures* |  |  |  | 0.0112 |
| *Always* | 7 (4.9) | 4<br>(57.14) | 3<br>(42.86) |  |
| *Sometime* | 98 (68.5) | 27<br>(27.55) | 71<br>(72.45) |  |
| *Never* | 38 (26.6) | 20<br>(52.63) | 18<br>(47.37) |  |
| *Engage in activities*<br>*with contaminated*<br>*water* |  |  |  | 0.5188 |
| *Daily* | 123 (86) | 44<br>(35.77) | 79<br>(64.23) |  |
| *Weekly* | 13 (9.1) | 6<br>(46.15) | 7<br>(53.8) |  |
| *Monthly* | 0 | 0 | 0 |  |
| *Rarely* | 5 (3.5) | 1<br>(20) | 4<br>(80) |  |
| *Never* | 2 (1.4) | 51<br>(35.66) | 92<br>(64.34) |  |
| *IL 4* | 57.1<br>(29.4, 312.5) | 47.6<br>(25.5, 259.3) | 66.7<br>(31.97, 316.55) | 0.2979 |
| *IL 10* | 26.7<br>(19.4, 50.9) | 33.53<br>(20.23, 50.58) | 26.27<br>(19.16, 51.22) | 0.7939 |
| *Eosinophil* | 0.12<br>(0.07, 0.4) | 0.11<br>(0.07, 0.28) | 0.12<br>(0.07, 0.49) | 0.3394 |
| *RBC* | 4.51<br>(4.12, 5) | 4.51<br>(4.12, 5.1) | 4.52<br>(4.115, 4.89) | 0.7013 |
| *Hb* | 12<br>(10.9, 13.2) | 12<br>(11.1, 13.3) | 11.95<br>(10.7, 13) | 0.8993 |
| *MCV* | 87<br>(81.5, 92) | 88 (81.5, 92) | 86.55 (81.5, 92) |  |
| *MCH* | 27.1<br>(24.4, 28.5) | 27.1<br>(24.8, 28.3) | 26.9 (24.2, 28.6) |  |
| *MCHC* | 30.1<br>(29.2, 30.9) | 30.2<br>(29.4, 30.9) | 30.1<br>(29.15, 30.9) |  |
| *Neutrophil* | 1.24<br>(1.01, 1.96) | 1.17<br>(0.93, 1.88) | 1.27<br>(1.01, 2.08) | 0.5551 |
| *Lymphocyte* | 2.4<br>(1.68, 2.94) | 2.1<br>(1.68, 2.7) | 2.45<br>(1.72, 3.4) | 0.0601 |
| *Monocyte* | 0.24<br>(0.14, 0.62) | 0.24<br>(0.14, 0.6) | 0.24<br>(0.14, 0.63) | 0.6128 |
| *Platelet* | 244<br>(190, 285) | 245<br>(180, 295) | 244<br>(191.5, 284.5) | 0.9027 |
| *PLR* | 104.9 (71.6, 140.4) | 108.7<br>(80.6, 142.3) | 97.6<br>(63.1, 138.5) | 0.2388 |
| *LMR* | 9.2<br>(4.2, 13.9) | 8.7<br>(3.9, 13.9) | 9.3<br>(5.0, 12.8) | 0.7535 |
| *NLR* | 0.58<br>(0.42, 0.86) | 0.63<br>(0.48, 0.83) | 0.56<br>(0.38, 0.89) | 0.2844 |

### Association in Logistic Regression

Table 2 presents the univariate analysis, showing the crude association between each variable and schistosomiasis, separately for males and females. For Males: Several factors were significantly associated with schistosomiasis. The strongest protective factor was access to clean water (OR=0.03, 95% CI: 0.00-0.29, p=0.0019), while the strongest risk factor was reporting any symptoms (OR=53.13, 95% CI: 6.08-464.48, p=0.0003). Using an unprotected water source was a major risk (OR=29.75, 95% CI: 3.51-252.33, p=0.0019), as was not using protective measures (OR=3.02, 95% CI: 1.04-8.79, p=0.0422). A higher lymphocyte count was also a significant risk factor (OR=2.11, 95% CI: 1.04-4.26, p=0.0378).

**Table 2.** Sex Differences in Correlates of Schistosomiasis in Univariate Analysis.

| Variable | Men |  | Women |  |
| --- | --- | --- | --- | --- |
|  | OR (95% CI) | P-value | OR (95% CI) | P-value |
| Age years | 0.98 (0.94, 1.02) | 0.3939 | 0.96 (0.93, 1.00) | **0.0264** |
| Access to clean water | 0.03 (0.00, 0.29) | **0.0019** | 0.19 (0.07, 0.52) | **0.0013** |
| Source of water for drinking | 29.75 (3.51, 252.33) | **0.0019** | 6.95 (2.37, 20.35) | **0.0004** |
| Do you use protective measures | 3.02 (1.04, 8.79) | **0.0422** | 7.46 (2.29, 24.29) | **0.0008** |
| Any symptoms related to Schistosoma | 53.13 (6.08, 464.48) | **0.0003** | 18.75 (6.51, 54.04) | **<0.0001** |
| IL-4 | 1.00 (1.00, 1.00) | 0.0590 | 1.01 (1.01, 1.01) | **<0.0001** |
| Eosinophil count | 10.37 (0.79, 136.56) | 0.0754 | 8.53 (2.30, 31.65) | **0.0014** |
| Hb | 0.96 (0.78, 1.18) | 0.7068 | 0.56 (0.41, 0.77) | **0.0003** |
| Mcv | 0.99 (0.94, 1.04) | 0.6397 | 0.94 (0.89, 0.99) | **0.0221** |
| Mch | 0.86 (0.72, 1.02) | 0.0835 | 0.80 (0.70, 0.91) | **0.0010** |
| MCHC | 0.78 (0.55, 1.10) | 0.1618 | 0.72 (0.55, 0.94) | **0.0143** |
| Lymphocyte count | 2.11 (1.04, 4.26) | **0.0378** | 2.27 (1.48, 3.48) | **0.0002** |
| Monocyte count | 2.68 (0.51, 14.00) | 0.2436 | 5.38 (1.80, 16.12) | **0.0026** |
| PLR | 1.00 (0.99, 1.01) | 0.6221 | 0.99 (0.98, 1.00) | **0.0063** |

For Females: Similar to males, access to clean water was protective (OR=0.19, 95% CI: 0.07-0.52, p=0.0013) and any symptoms were a strong risk factor (OR=18.75, 95% CI: 6.51-54.04, p<0.0001). Numerous other variables were significant for females that were not for males. These included older age (OR=0.96, p=0.0264), lower red blood cell indices (hb, mcv, mch, mchc), higher IL-4 (OR=1.01, p<0.0001), higher eosinophil count (OR=8.53, p=0.0014), and higher monocyte count (OR=5.38, p=0.0026). A lower Platelet-to-Lymphocyte Ratio (PLR) was also associated with infection (OR=0.99, p=0.0063).

Table 3 presents the results of the multivariate analysis, which identifies the independent correlates of schistosomiasis after adjusting for other variables in the model.

**Table 3.**
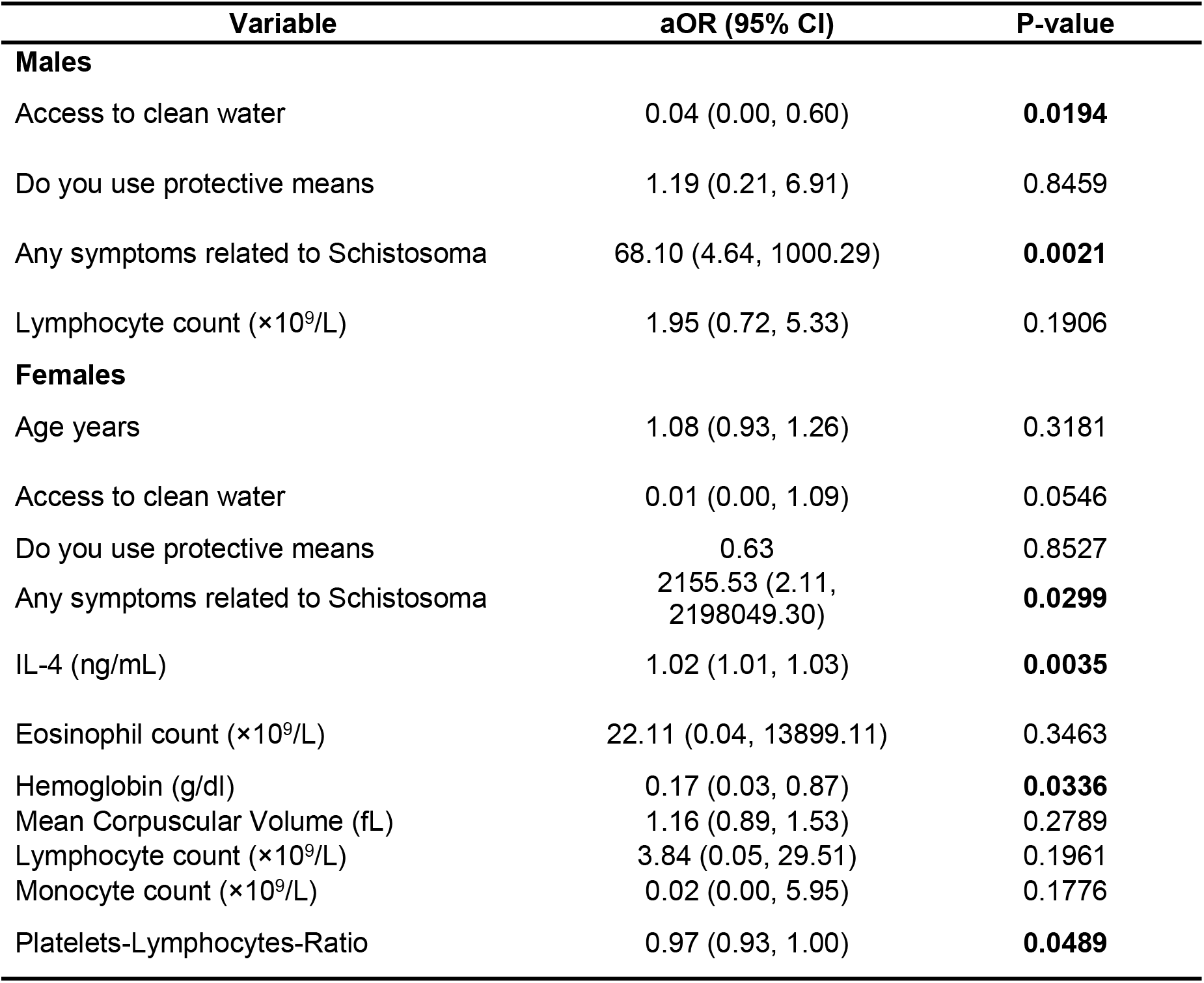
Sex Differences in Correlates of Schistosomiasis in Multivariate Analysis.

For Males: When controlling for other factors, only two variables remained significant independent correlates. Access to clean water remained a significant protective factor (OR=0.04, 95% CI: 0.00-0.60, p=0.0194), and reporting any symptoms remained significantly associated with schistosomiasis infection (OR=68.10, 95% CI: 4.64-1000.29, p=0.0021). The use of protective measures and lymphocyte count, which were significant in the univariate analysis, were no longer significant in the multivariate model.

For Females: The multivariate model for females was more complex. The most prominent independent significant factor was reporting any symptoms (OR=2155.53, 95% CI: 2.11-2198049.30, p=0.0299), with an extremely wide confidence interval. Higher levels of IL-4 were a significantly associated with Schistosomiasis (OR=1.02, 95% CI: 1.01-1.03, p=0.0035), while lower hemoglobin (hb) was a significant protective factor (OR=0.17, 95% CI: 0.03-0.87, p=0.0336). A lower Platelet-to-Lymphocyte Ratio (PLR) also remained a significant independent correlate (OR=0.97, 95% CI: 0.93-1.00, p=0.0489). Access to clean water and other variables like age and eosinophil count were not significant in the adjusted model.

## Discussion

The results of this study paint a compelling picture of distinct disease experiences between males and females in this endemic area. The univariate analysis suggests that while both sexes are vulnerable to infection through water exposure, the physiological consequences differ markedly. The female cohort exhibited a stronger and more diverse immune and hematological response, characterized by a Th2-polarized immune signature (elevated IL-4), eosinophilia, and significant anemia. This pattern suggests that females may mount a more vigorous, though ultimately ineffective, immune reaction to the parasite, leading to greater hematological compromise.

This finding aligns with the established paradigm of sex-based immunology, where females often mount more robust innate and adaptive immune responses to a variety of pathogens, including helminths [11]. The enhanced Th2 profile observed in women, characterized by elevated IL-4, is consistent with studies showing that estradiol can potentiate Th2-type responses [24]. Furthermore, the pronounced eosinophilia in females supports this, as eosinophils are key effector cells in the Th2-driven anti-helminth response [25]. However, this heightened immunity appears to come at a cost. The significant anemia observed in infected women is a critical finding, reinforcing the link between a potent inflammatory response and hematological dysregulation. Chronic schistosomiasis is a known driver of anemia of inflammation (AI), where hepcidin-mediated iron sequestration and the direct effects of inflammatory cytokines like IL-6 on erythropoiesis lead to reduced hemoglobin [26]. Our data suggest that the stronger Th2-inflammatory milieu in females may exacerbate this process, making them particularly vulnerable to this debilitating comorbidity [27].

The multivariate analysis further sharpens this distinction. For males, the model is straightforward, pointing directly to environmental risk (lack of clean water) and clinical presentation (symptoms). This implies that for males, infection is primarily a function of exposure and acute disease, a pattern frequently reported in epidemiological studies that attribute higher male prevalence to occupational and recreational water contact [18]. For females, the model is more complex and biologically nuanced. The persistence of IL-4 as a strong independent correlate confirms the central role of a Th2 response in the female pathophysiology of schistosomiasis, beyond what is explained by exposure alone [28]. Furthermore, anemia (low hemoglobin) remains a key feature even after adjusting for other factors, highlighting it as a major clinical outcome for infected women. This is of particular public health importance given the synergistic effects of anemia and female genital schistosomiasis (FGS) on maternal and reproductive health outcomes [3,20].

The significance of the platelet-to-lymphocyte ratio (PLR), a potential marker of inflammation, adds another layer to the female-specific pathological profile. An elevated PLR has emerged as a biomarker signifying systemic inflammation in various chronic infectious and non-infectious diseases [29]. In the context of schistosomiasis, it may reflect the complex interplay between a chronic granulomatous response to eggs (involving platelet-activating factors) and the subsequent immunoregulatory phase that modulates lymphocyte activity [30]. Its prominence in the female model suggests a more pronounced inflammatory state or a distinct immunopathological process compared to males, warranting further investigation into the role of platelets and coagulation in female-specific schistosomiasis morbidity [31].

The immense odds ratio for symptoms in the female multivariate model, with an extremely wide confidence interval, should be interpreted with caution. It indicates an exceptionally strong association but also reflects the model’s instability, likely due to the high correlation between symptoms (like hematuria) and active infection in this dataset, a well-documented phenomenon in S. haematobium studies [32]. This collinearity makes it statistically challenging to disentangle the independent effect of symptoms from the infection itself in this specific cohort.

## Conclusion

In conclusion, this study demonstrates that urogenital schistosomiasis is not a uniform disease across sexes. Males appear to be primarily affected by external, exposure-related factors. In contrast, females experience the infection with a distinct immunopathological phenotype, involving a pronounced Th2 immune response and significant anemia. These findings move beyond a one-size-fits-all understanding of schistosomiasis and resonate with calls for a more nuanced, pathophysiologically informed approach to NTDs [33]. They advocate for gender-sensitive strategies, where clinical management for women closely monitors for anemia and immunological sequelae, and public health interventions are tailored to address the specific risk profiles and disease manifestations of each sex [34]. Future research should focus on longitudinal studies to determine if these sex-specific correlates predict differential reinfection rates post-treatment and on mechanistic studies to unravel the hormonal and genetic drivers of this immunological dimorphism.

## Data Availability

The raw data underlying the results is available in the supporting information S2.

## KEY POINTS

- **Distinct Sex-Specific Profiles:** The study identifies fundamentally different correlates of *S. haematobium* infection between sexes. Male infection is primarily linked to environmental exposure (lack of clean water), while female infection is characterized by a pronounced immunological (elevated IL-4) and hematological (anemia) response.
- **Female-Specific Immune-Hematological Link:** In females, infection is independently associated with a stronger Th2-polarized immune response (higher IL-4) and significant anemia (lower hemoglobin), suggesting a more vigorous immunopathological reaction that contributes to key morbidity.
- **Implications for Differential Management:** The findings underscore the necessity for sex-differentiated approaches in both clinical assessment, where women may require closer monitoring for anemia, and in public health control strategies that address distinct risk profiles.

## Availability of data and materials

The raw data underlying the results is available in the supporting information S2.

S1 Strobe

S1. Data

